# Exploratory factor analysis as a validation tool in a questionnaire to evaluate the uptake of cataract surgery

**DOI:** 10.1101/2025.01.29.25321326

**Authors:** Mohamad Aziz Salowi, Nyi Nyi Naing, Hajar Ahmad Rosdi, Norasyikin Mustafa, Wan Radziah Wan Nawang, Siti Nurhuda Sharudin, Nor Anita Che Omar, Azlina Mokhtar

## Abstract

**Background:** Exploratory Factor Analysis (EFA) evaluates construct validity by identifying underlying relationships among questionnaire items, helping determine the number of latent constructs. This study aimed to validate a newly developed questionnaire on cataract surgery uptake among the community by interviewing subjects aged 50 years and older.

**Methods:** A bilingual questionnaire, based on theoretical factors of knowledge, perception, attitude, and practice, was developed through literature review and expert interviews. It underwent forward-backward translation, content validity, and face validity assessment, followed by discussions with panels of eye care experts. Data for EFA were collected through interviews with subjects aged 50 and older, who had visual acuity worse than 6/18 due to cataracts or had undergone cataract surgery. Responses were analyzed for factor loadings and reliability. Principal Axis Factoring (PAF) was used for factor extraction, initially based on Eigenvalues > 1, and later fixed at eight factors. Factors were rotated using Varimax with Kaiser Normalization. Items with weak factor loadings were removed iteratively, with re-analysis conducted after each removal.

**Results:** A total of 287 subjects were recruited and consented to EFA. All 40 items in the questionnaire (knowledge, attitude, perception and practice) were combined and analysed. The Kaiser-Meyer-Olkin (KMO) test was 0.714, indicating that factor analysis was suitable for exploring the underlying structure. Bartlett’s test of Sphericity was highly significant (*p* < 0.001), indicating that the variables (questions) were sufficiently interrelated for factor analysis. Factors 3 and 7 had only two questions, although they showed fairly good loading with satisfactory correlation. The Cronbach’s alpha reliability of all the factors varies from 0.50 to 0.72 (total = 0.72). The final questionnaire consisted of six factors and 22 questions.

**Conclusions:** The developed and validated questionnaire demonstrated an acceptable factor structure related to knowledge, attitude, perception, and practice in evaluating cataract surgery uptake. This tool provides a reliable method for assessing cataract surgery uptake in the community.

## Introduction

Malaysia is divided into six administrative regions for the Prevention of Blindness and Low Vision (PBL) eye care service monitoring: Northern, Eastern, Central, Southern, Sabah, and Sarawak (Fig 1). Six simultaneous population surveys on blindness were done separately in 2014 in each region to generate evidence on the magnitude and causes of vision impairment as proposed in the World Health Organization (WHO) Global Action Plan 2014-2019 [1,2].

**Fig 1.**
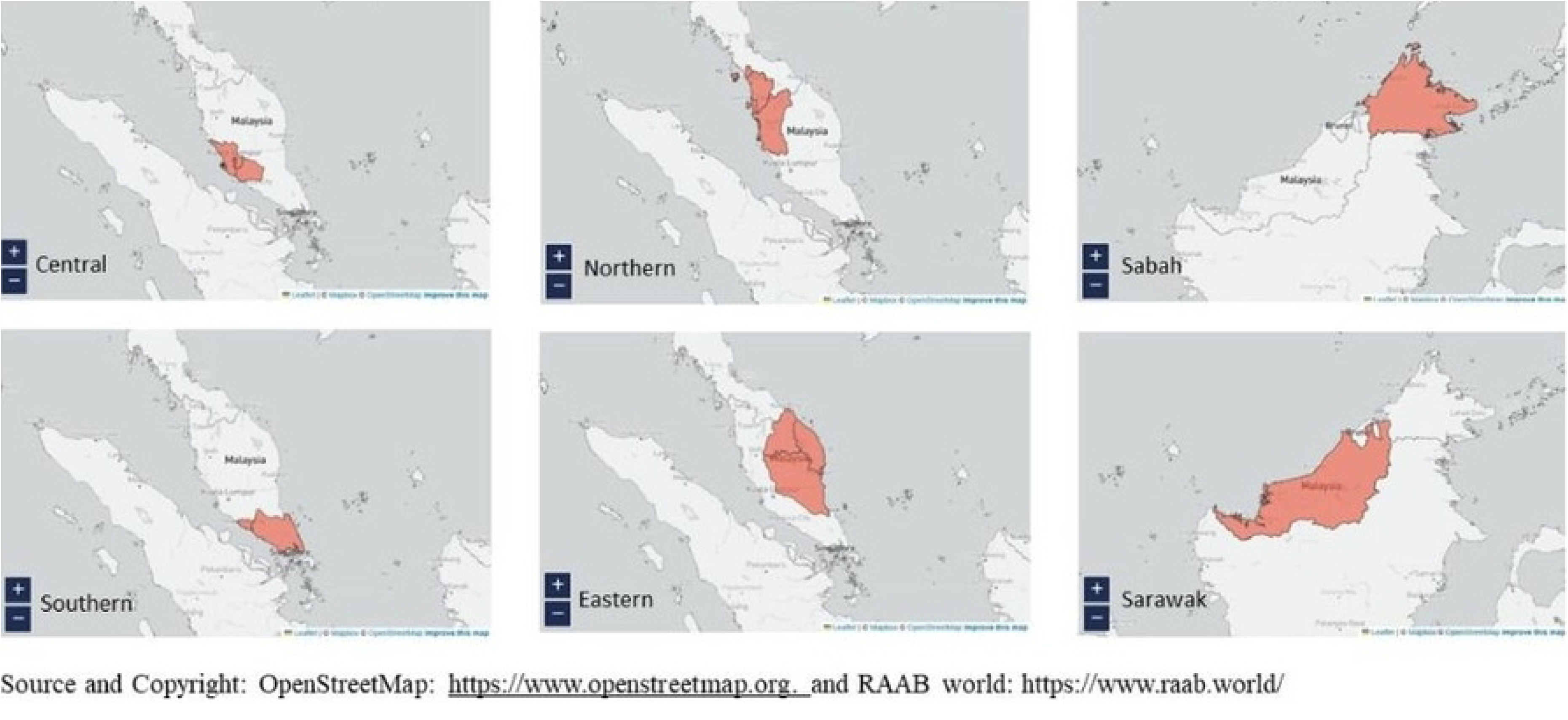
Administrative regions for the Prevention of Blindness and Low Vision (PBL) eye care service monitoring in Malaysia.

Reprinted from [Rapid Assessment of Avoidable Blindness: Survey Data. Available from: https://www.raab.world/survey-dataRAAB world] under a CC BY license, with the permission from OpenStreetMap Foundation (OSMF), original copyright year 2024

The core findings of the surveys (National Eye Survey, NES II) were a higher prevalence of blindness and cataract blindness in the peripheral regions, namely Sabah, Sarawak, Northern, and Eastern Regions, and significant disparities in cataract surgery outcomes between all the regions [3]. As part of the post-NES II survey action plan, the Cataract Clinic Ministry of Health Malaysia *(Klinik Katarak, Kementerian Kesihatan Malaysia*, *KK-KKM*) was piloted in Sarawak and the Eastern Region to address the cataract blindness issue (Fig 2). However, after more than 10 years of operation, the need for evidence-based information to evaluate and improve its implementation, including its service delivery and uptake by the population, became increasingly urgent and important. The objective, concept, and work process of *KK-KKM* were endorsed by the WHO when it was selected as a Case Study for the Western Pacific WHO Innovation Challenge in 2021/2022 [4]. The service brands had been established, and specific funds were regularly allocated to the projects. However, the population-based data on blindness and hospital-based data on cataract surgery differed between both regions; a lower percentage of phacoemulsification in the Eastern Region despite less percentage of patients presented with advanced vision (hard cataract) and generally higher yearly output and lower complication percentage in Sarawak [5–7]. We postulate that these data disparities could result from community-related factors and their interactions, which we would like to identify and rectify, hence the need for a questionnaire as an instrument to collect data from the population within the regions.

**Fig 2.**
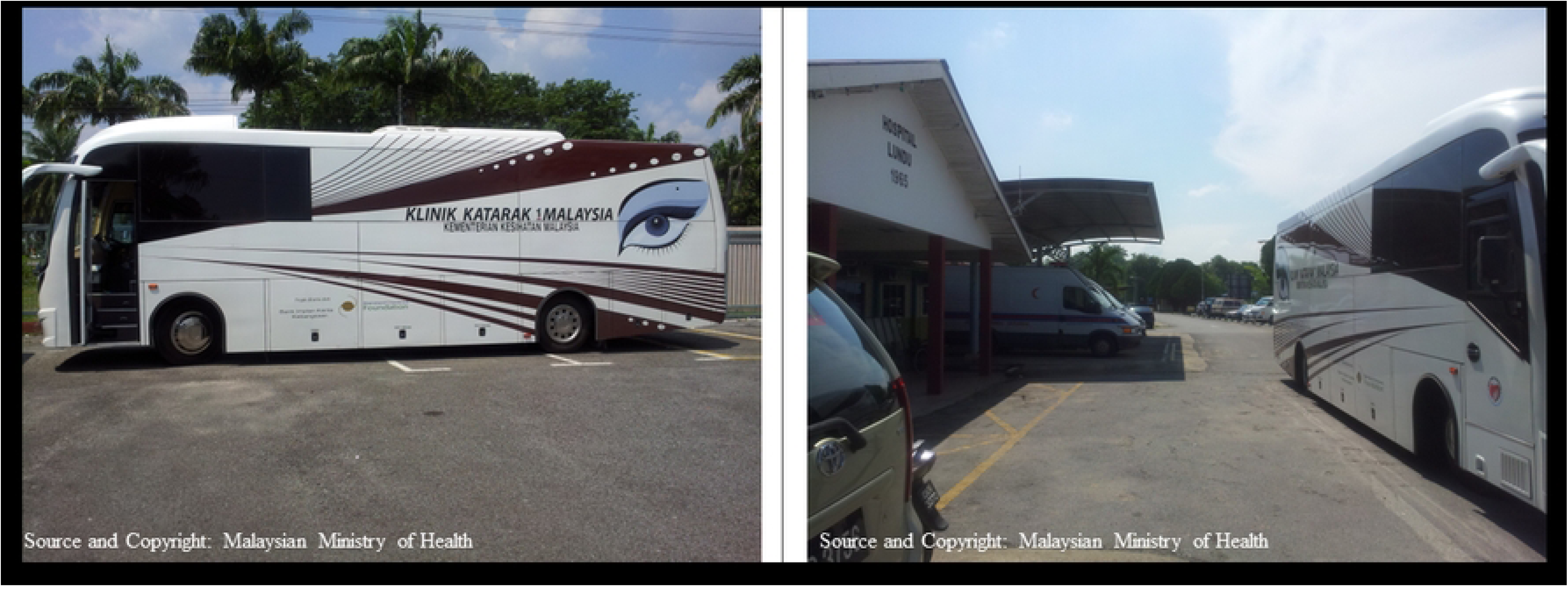
The mobile unit parked in front of a provincial hospital in Sarawak.

## Methods

A literature study was conducted to search for and identify existing questionnaires, relevant items, and scales for available resources on knowledge, attitude, and practice related to eye care evaluation. Discussion by a panel of eye care experts (comprised of public health ophthalmologists, clinical ophthalmologists, optometrists, biostatisticians, nurses, and educators) and interviews of the eye care providers on the ground were conducted to qualitatively explore the factors influencing the uptake of services by the community. The questionnaire was developed simultaneously in English and Malay (the official national language) and underwent twice forward-backwards translations by two independent Ophthalmologists. Content validity was done by a panel consisting of a vitreoretinal surgeon, a comprehensive ophthalmologist, a paediatric optometrist, a community optometrist, a government pensioner, and a businessman who was a Rotary Club member. Face validity interviews were conducted on ten patients aged 50 years and older attending an eye clinic (for various reasons) at one of the hospitals in the Northern Region of Peninsular Malaysia. The content validity index (CVI) and face validity index (FVI) were discussed separately in another manuscript. A preliminary 51-item questionnaire developed through the initial process was rephrased and reduced to a 40-item questionnaire based on CVI and FVI results and discussion with the panel experts. This questionnaire was used to collect data for exploratory factor analysis (EFA) and reliability evaluation.

EFA and reliability data collection was conducted from 1st September 2020 to 31st October 2020. The sample size required was 5-10 subjects per questionnaire item [8]. As the number of items was 40, the sample size estimated was 200-400 participants. Six interviewers were hired for data collection purposes. They underwent three training days, including lectures on basic eye anatomy and selected eye diseases, especially cataracts. They were also given hands-on practical sessions on eye examination, visual acuity assessment and staff interviews using the questionnaire.

The questionnaire was developed to identify factors influencing the uptake of cataract surgery within the community nationwide, but during COVID-19, data collectors were only permitted to interview subjects within the selected hospital compounds due to the implementation of the Movement Control Order (MCO) [9]. Subjects in this study, therefore, were patients or their relatives who attended the various Outpatient Departments within the hospital facilities in mixed urban-rural settings. Similarly, due to the MCO, hospitals and health facilities were also selected, subject to approval from the Health State Offices. It was done in Sarawak (one urban and three provincial hospitals), Eastern Region (one urban and eight provincial hospitals), Northern Region (one community health clinic) and Central Region (one community health clinic). The interviews followed the updated Standard Operating Procedure (SOP) for COVID-19 (mainly physical distancing, wearing masks, and washing hands after each interview). The subjects were briefed about the study, the benefits, the data used, and the expected study outcome. Visual acuity was measured using Vis-Screen, a validated mobile application developed for easy and rapid identification of vision impairment by any layperson [10,11]. All participants provided written informed consent, as detailed in the ethics statement. The consent process included a clear explanation of the study’s purpose, procedures, potential risks, and participant rights, in a language accessible to all. Participants were given ample time to review the form and ask questions before consenting. The questionnaire was then administered via an interview conducted by the data collectors, and the scales were documented accordingly.

### Statistical Analysis

Statistical analysis was performed using Statistical Package for Social Science (SPSS), version 26.0 (SPSS, Inc., Chicago, III., USA) for Windows. EFA was conducted to assess the construct validity of all the domains/factors. Factors were extracted using the Principal Axis Factoring method. Rotation was done using Varimax with Kaiser Normalization. Kaiser-Meyer-Olkin’s was measured to assess sampling adequacy (KMO) and Bartlett’s test of the Sphericity was done to determine if there was redundancy between the variables. Components with Eigenvalues of greater than one (>1) were retained as components using parallel analysis and scree plots. Items with a loading factor of more than plus or minus 0.3 were considered acceptable [12]. The items’ internal consistency (IC) was measured using Cronbach’s Alpha Coefficient. The questionnaire items were supposed to represent a measure of good internal consistency if the total Cronbach’s alpha value was more than 0.7 [13]. The finalised questionnaire was later evaluated for fitness using confirmatory factor analysis (CFA) and used to collect data for the population surveys for Sarawak and the Eastern Region. The manuscript for CFA and the population surveys were discussed in another manuscript.

## Results

Following content validity, face validity, forward-backwards translations and discussion among the study committee members, four theoretical factors with 40 questionnaire items were developed. Knowledge had three response choices (1= Yes, 2= Unsure, 3= No). Attitude and Perception had five response choices (1= Strongly Agree 2= Agree 3= Unsure 4= Not Agree 5= Strongly Not Agree), and Practice had four response choices (1= Always 2= Often 3= Seldom 4= Never) (Table 1).

**Table 1.**
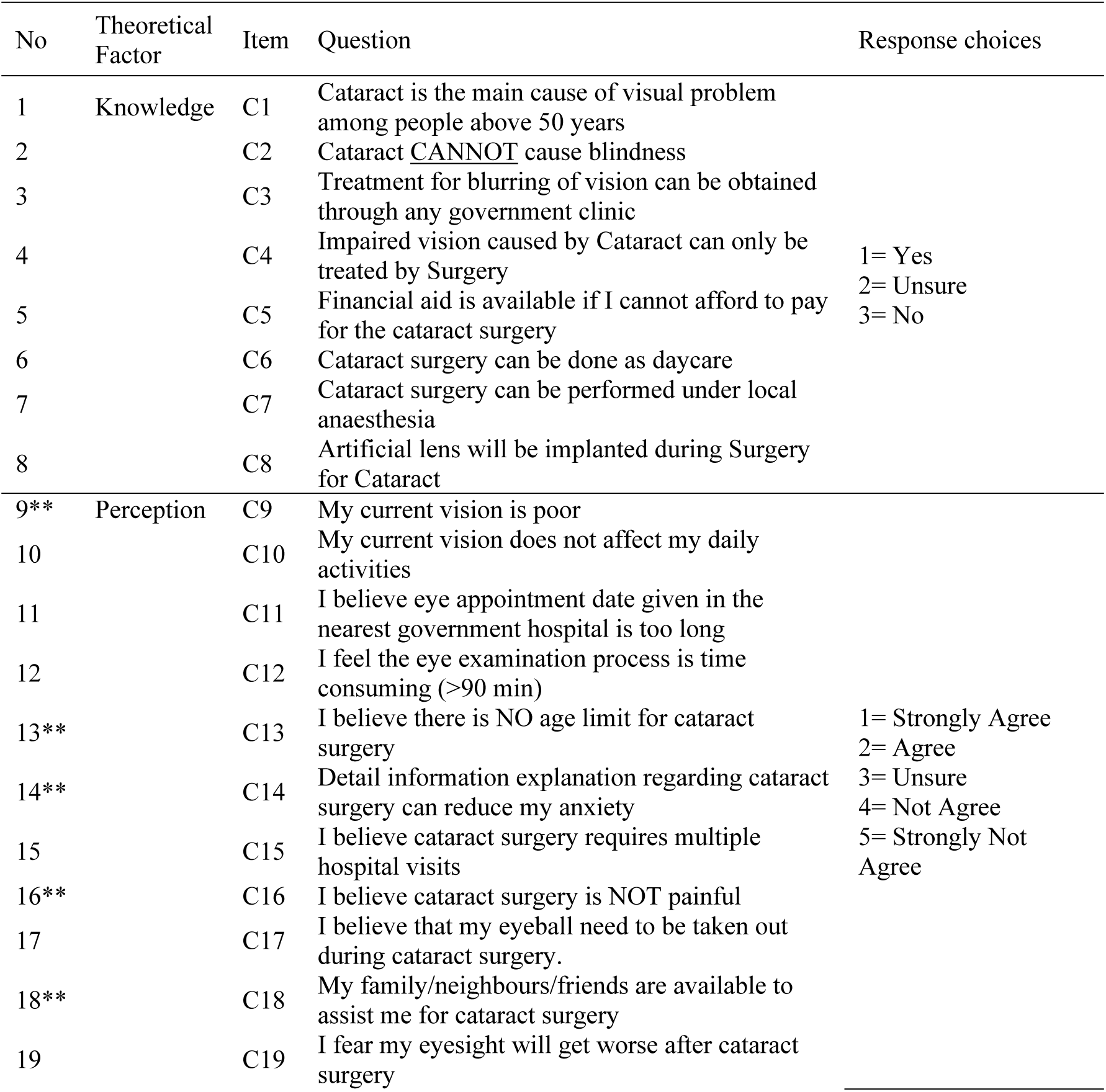
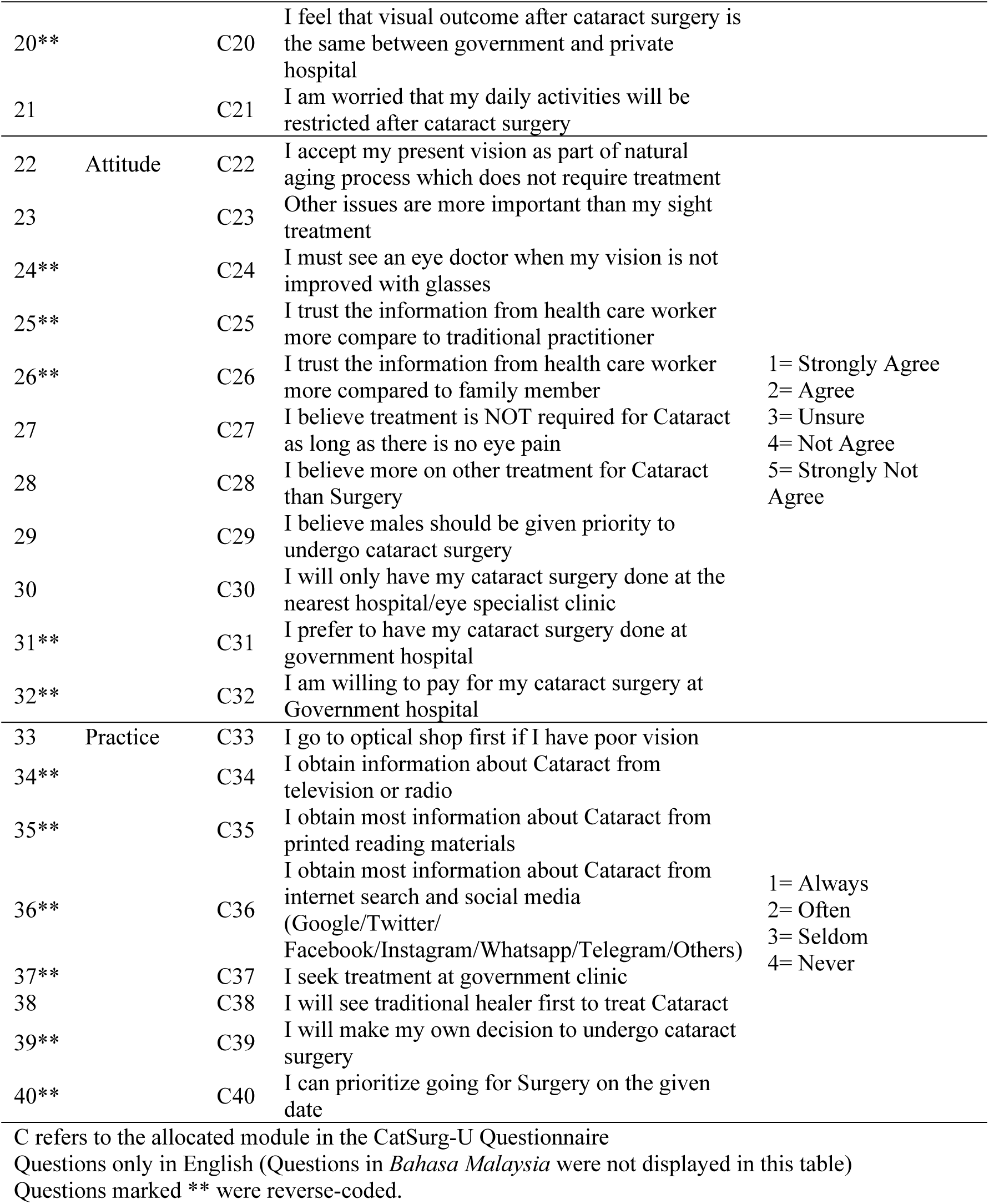
Questionnaire Items and Theoretical Factors (40 Items)

A total of 287 subjects were recruited and consented to exploratory factor analysis (EFA) data collection. All 40 items in the questionnaire (knowledge, attitude, perception, and practice) were combined and analysed for possible interaction. The Kaiser-Meyer-Olkin (KMO) test was 0.71, indicating that factor analysis was suitable for exploring the underlying structure. Bartlett’s test of Sphericity was highly significant (*P*< 0.001), indicating that the variables (questions) were sufficiently interrelated for factor analysis (Table 2). The factors were analysed using the Principal Axis Factoring (PAF) method, preliminarily extracted based on Eigenvalues >1, and then later based on fixing the number of factors to eight. They were rotated using Varimax with the Kaiser Normalization method. Removal of items with weak factor loading was done one item at a time, followed by re-analysis and item removal. Factors were renamed: Factor 1=Attitude towards Treatment, Factor 2=Practice towards Information, Factor 3=Attitude towards Information, Factor 4=Perception to Own Sight, Factor 5=Knowledge on Surgery, Factor 6=Practice on Surgery, Factor 7=Perception on Outcome, and Factor 8=Knowledge on Cataract (Table 2).

**Table 2.** Results of the Exploratory Factor Analysis and Reliability test (from 40 items, reduced to 26 items)

| No | Factor | Item | Mean | SD | Communalities | Factor loading | Corrected ITC | Alpha |
| --- | --- | --- | --- | --- | --- | --- | --- | --- |
| 1 | 1 | C18 | 4.07 | 0.84 | 0.35 | 0.44 | 0.43 | 0.69 |
| 2 | (Attitude towards Treatment) | C23 | 3.69 | 1.01 | 0.38 | 0.41 | 0.43 |  |
| 3 |  | C28 | 3.67 | 0.90 | 0.38 | 0.41 | 0.37 |  |
| 4 |  | C29 | 3.79 | 0.99 | 0.38 | 0.55 | 0.47 |  |
| 5 |  | C31 | 4.17 | 0.71 | 0.36 | 0.57 | 0.46 |  |
| 6 |  | C37 | 3.25 | 0.81 | 0.41 | 0.60 | 0.38 |  |
| 7 | 2 | C34 | 1.65 | 0.76 | 0.49 | 0.62 | 0.51 | 0.72 |
| 8 | (Practice towards Information) | C35 | 1.56 | 0.78 | 0.63 | 0.76 | 0.57 |  |
| 9 |  | C36 | 1.57 | 0.81 | 0.46 | 0.66 | 0.53 |  |
| 10 | 3 | C25 | 4.27 | 0.66 | 0.92 | 0.89 | 0.71 | 0.83 |
| 11 | (Attitude towards Information) | C26 | 4.17 | 0.66 | 0.55 | 0.70 | 0.71 |  |
| 12 | 4 | C9 | 3.62 | 1.01 | 0.53 | 0.64 | 0.40 | 0.60 |
| 13 | (Perception to Own Sight) | C10 | 2.98 | 1.20 | 0.36 | 0.54 | 0.34 |  |
| 14 |  | C22 | 3.14 | 1.11 | 0.41 | 0.50 | 0.45 |  |
| 15 |  | C27 | 3.34 | 1.08 | 0.55 | 0.46 | 0.34 |  |
| 16 | 5 | C6 | 1.47 | 0.68 | 0.33 | 0.44 | 0.36 | 0.60 |
| 17 | (Knowledge on Surgery) | C7 | 1.49 | 0.59 | 0.63 | 0.78 | 0.53 |  |
| 18 |  | C8 | 0.87 | 0.99 | 0.37 | 0.51 | 0.41 |  |
| 19 | 6 | C32 | 3.27 | 1.15 | 0.48 | 0.66 | 0.39 | 0.61 |
| 20 | (Practice on Surgery) | C39 | 2.43 | 1.17 | 0.45 | 0.56 | 0.52 |  |
| 21 |  | C40 | 3.20 | 0.91 | 0.43 | 0.34 | 0.37 |  |
| 22 | 7 | C21 | 2.71 | 0.99 | 0.39 | 0.60 | 0.31 | 0.48 |
| 23 | (Perception on Outcome) | C19 | 2.91 | 1.06 | 0.22 | 0.45 | 0.31 |  |
| 24 | 8 | C1 | 1.42 | 0.91 | 0.26 | 0.35 | 0.33 | 0.50 |
| 25 | (Knowledge on Cataract) | C3 | 1.73 | 0.54 | 0.24 | 0.47 | 0.32 |  |
| 26 |  | C5 | 1.43 | 0.66 | 0.47 | 0.65 | 0.36 |  |
| Kaiser-Meyer-Olkin Measure of Sampling Adequacy (KMO): |  |  | 0.71 |  |  |  |  |  |
| Bartlett's Test of Sphericity: |  | Approx. Chi-Square: | 1850.107 |  |  |  |  |  |
|  |  | df: | 325 |  |  |  |  |  |
|  |  | P-value: | <0.001 |  |  |  |  |  |

Questions only in English; SD=Standard Deviation; ITC=Item-Total Correlation Method: Rotated Factor Matrix with Eigenvalue >1 Extraction: Principal Axis Factoring. Rotation: Varimax with Kaiser Normalization. Total Alpha = 0.72 (range 0.50 to 0.72)

Factor 3 (C25:*I trust the information from health care worker more compare to traditional practitioner*, C26: *I trust the information from health care worker more compared to family member*) and Factor 7 (C19: *I fear my eyesight will get worse after cataract surgery*, C21: *I am worried that my daily activities will be restricted after cataract surger*y) were removed because only two items were loaded into one factor. Cronbach’s alpha reliability of all the factors varies from 0.50 to 0.72 (Table 2). Total Cronbach’s Alpha for the final model (22 items) was 0.72. There were two questions in Factor 1, two questions in Factor 4, and one question in Factor 6, which loaded into the respective factors instead of loading into the intended theoretical factors; Factor 1 = Attitude towards Treatment (C18 was theoretically Perception, C37 was theoretically Practice), Factor 4=Perception to own Sight (C22 and C27 were theoretically Attitude) and Factor 6=Practice on Surgery (C32 was theoretically Attitude) (Tables 1 and Table 2).

Although most of the questions loaded satisfactorily into the respective factors (except for C1 and C40), the values of communalities were mostly low (ranging from 0.22 to 0.92). The Corrected Item-Total Correlation (ITC) values were all above 0.3 (Table 3). The final questionnaire comprised six factors and 22 questions.

**Table 3.**
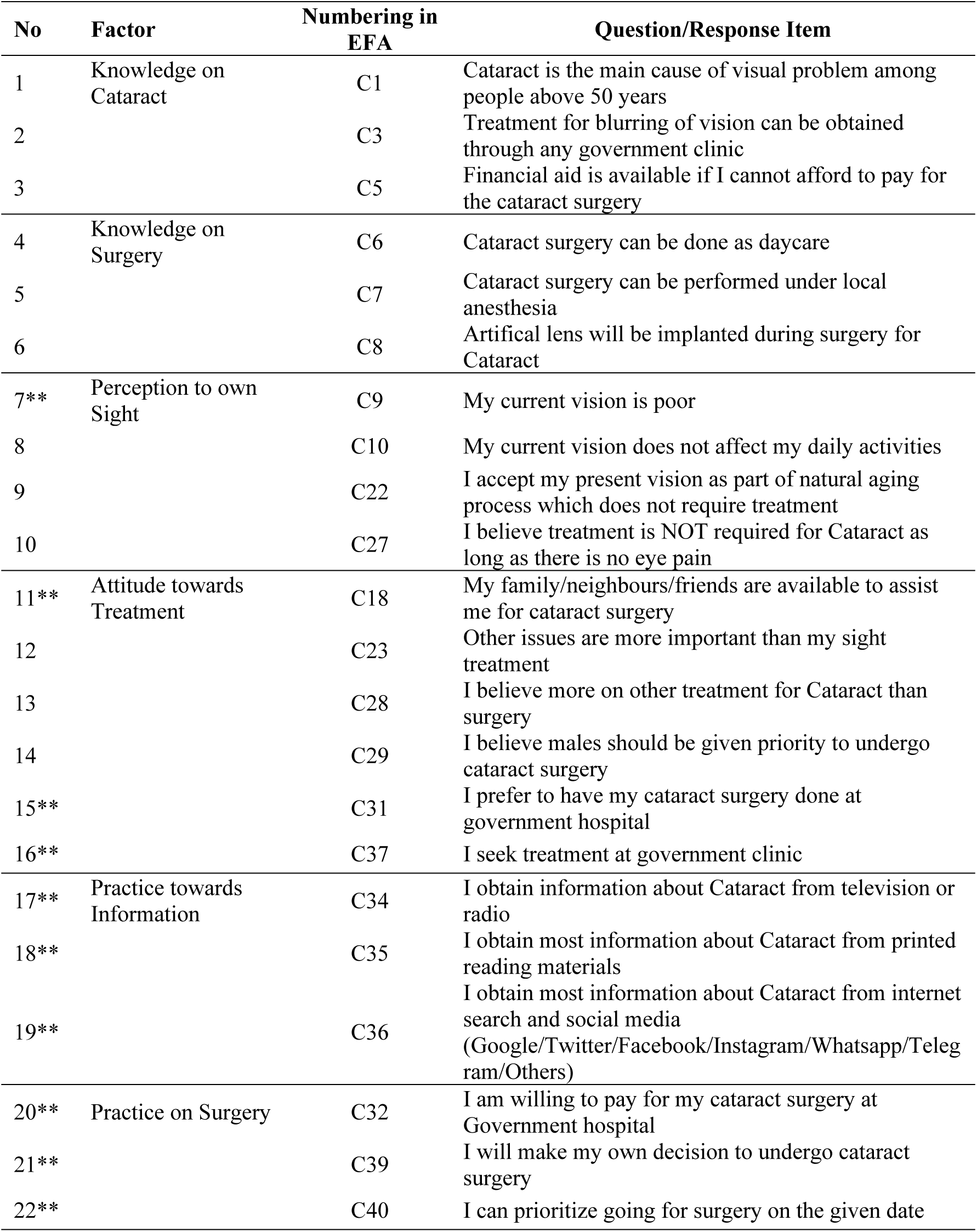
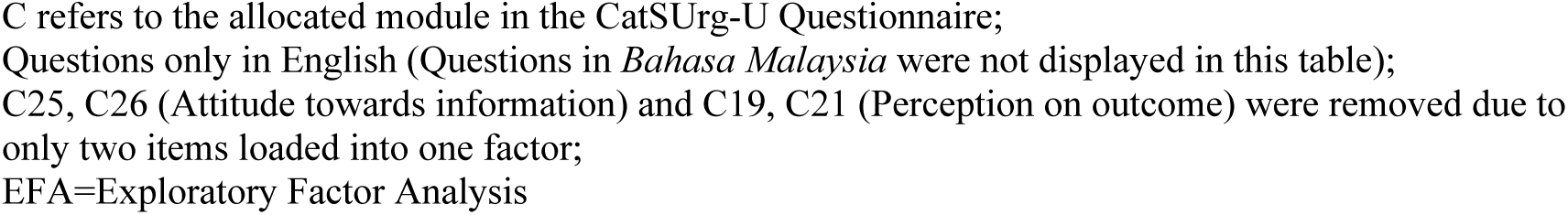
Final Factors and Questionnaire Items (22 Items)

| No | Factor | Numbering in EFA | Question/Response Item |
| --- | --- | --- | --- |
| 1 | Knowledge on Cataract | C1 | Cataract is the main cause of visual problem among people above 50 years |
| 2 |  | C3 | Treatment for blurring of vision can be obtained through any government clinic |
| 3 |  | C5 | Financial aid is available if I cannot afford to pay for the cataract surgery |
| 4 | Knowledge on Surgery | C6 | Cataract surgery can be done as daycare |
| 5 |  | C7 | Cataract surgery can be performed under local anesthesia |
| 6 |  | C8 | Artificial lens will be implanted during surgery for Cataract |
| 7** | Perception to own Sight | C9 | My current vision is poor |
| 8 |  | C10 | My current vision does not affect my daily activities |
| 9 |  | C22 | I accept my present vision as part of natural aging process which does not require treatment |
| 10 |  | C27 | I believe treatment is NOT required for Cataract as long as there is no eye pain |
| 11** | Attitude towards Treatment | C18 | My family/neighbours/friends are available to assist me for cataract surgery |
| 12 |  | C23 | Other issues are more important than my sight treatment |
| 13 |  | C28 | I believe more on other treatment for Cataract than surgery |
| 14 |  | C29 | I believe males should be given priority to undergo cataract surgery |
| 15** |  | C31 | I prefer to have my cataract surgery done at government hospital |
| 16** |  | C37 | I seek treatment at government clinic |
| 17** | Practice towards Information | C34 | I obtain information about Cataract from television or radio |
| 18** |  | C35 | I obtain most information about Cataract from printed reading materials |
| 19** |  | C36 | I obtain most information about Cataract from internet search and social media (Google/Twitter/Facebook/Instagram/Whatsapp/Telegram/Others) |
| 20** | Practice on Surgery | C32 | I am willing to pay for my cataract surgery at Government hospital |
| 21** |  | C39 | I will make my own decision to undergo cataract surgery |
| 22** |  | C40 | I can prioritize going for surgery on the given date |

C refers to the allocated module in the CatSurg-U Questionnaire; Questions only in English (Questions in *Bahasa Malaysia* were not displayed in this table); C25, C26 (Attitude towards information) and C19, C21 (Perception on outcome) were removed due to only two items loaded into one factor; EFA=Exploratory Factor Analysis

## Discussion

A questionnaire is a research tool consisting of a series of questions designed to gather information from respondents. The recommended and available questionnaire for gap analysis in eye care services is the Eye Care Services Assessment Tool (ECSAT), developed by the WHO and released to member states in 2013 and 2022. ECSAT is designed to evaluate service and is used for eye care planning. The questionnaire responses grade the eye health system in the countries according to maturity level by scores [14]. However, it only receives and analyses responses from the policymakers rather than the people who utilise the services on the ground, and therefore, it is inappropriate for this study’s purposes. In eye care, most other questionnaires are more commonly used in hospital settings for clinical purposes or service quality assessment [15–19]. To the best of our knowledge, this is the first study in the literature that describes the development of a validated eye care questionnaire for community evaluation with reasonably acceptable construct validity and reliability. It examines the factors influencing the uptake of cataract surgical services using objective assessment of the respondents and eye examination (lens status). The questionnaire intended to help health policymakers identify the factors in the community that might have caused the outcome service discrepancy between Sarawak and the Eastern Region. The difference occurred despite the actively running mobile community cataract outreach program operating cataracts at the district hospitals in remote areas within both regions.

This study described the chronological process of designing, developing, and validating a KAP (Knowledge, Attitude and Practice) questionnaire to determine latent factors with corresponding response items that could represent the factors affecting the uptake of cataract surgical services [20]. The process commenced with 51 questions following brainstorming sessions, consultations, and discussions with multilevel community-based eye care providers. The content validity index (CVI), face validity index (FVI) and a series of discussions with the research committee and other stakeholders reduced the number of questions to 40 items. At this stage, most questions were rephrased based on the responses received during the Content and Face Validity exercises. The aim was to make them more engaging, easier to understand and suitable for laypeople [21–23].

The Principal Axis Factoring (PAF) method was chosen over the Principal Component Analysis (PCA) to extract factors from the 40 questionnaire items. This decision was made to focus more on the common variance shared among variables rather than the total variance, ensuring a more solid and refined understanding of the data. The dataset was thoroughly explored in PAF to identify latent constructs and simplify or reduce the number of response items/questions. The Varimax technique with Kaiser normalization was applied simultaneously to simplify the interpretation of factor loadings. This method ensured that the clear demarcation values between high and low loadings would help make the factors interpretable [24–28].

The Kaiser-Meyer-Olkin (KMO) test was done to determine whether the dataset was adequate for factor analysis. In this study, the value was 0.714, indicating that factor analysis was suitable for exploring the underlying structure. Bartlett’s test of Sphericity was used to assess the null hypothesis (that there were no correlations between the variables). If there were no correlations between variables (questions), then factor analysis would not be appropriate. In this study, the correlation was large (approximate Chi-square =1850.107), and the *P*-value was highly significant (*P*-value <0.001); therefore, the null hypothesis was rejected, indicating that the variables (questions) were sufficiently interrelated for factor analysis.

Communalities were used to assess how well the extracted factors represented the variables (questions); low communalities in this study (ranging from 0.22 to 0.92) might indicate that the factors did not adequately explain the variables (questions). A rule of thumb is that communalities should ideally be higher than 0.5 or 0.6 to suggest that the factors extracted adequately explain a significant portion of the variance in the variables. However, it’s important to consider the values by looking at other factors, too, such as factor loadings and theoretical considerations about the underlying structure of the data.

All the domains were explored in combination (including the domain on “knowledge”) using EFA, not as individual domains. Analysing all the domains with resulting good factor loadings has been reported by other authors [29–31]. From a total of eight questions on “Knowledge”, two questions were removed due to poor factor loading: C2 = *Cataract CANNOT cause blindness,* and C4 = *Impaired vision caused by Cataract can only be treated by Surgery* (both factor loading<0.3). Combining all the domains enabled a holistic exploration of all latent constructs per the study’s conceptual framework, which suggested potential interrelationships /interactions between the domains in evaluating cataract surgery uptake in the community. For example, knowledge on cataract or knowledge on surgery might influence the individual’s attitude towards surgery (having knowledge might reduce anxiety before surgery).

On the reliability assessment, Cronbach’s Alpha, which measured internal consistency reliability, ranged from 0.50 to 0.72 (total = 0.72), suggesting that each factor’s variables (questions) moderately correlated with each other and reliably measured a single underlying construct. All items had corrected item-total correlations (ITC) above 0.3, indicating a moderate correlation between the individual item and the scale’s total score. Both Cronbach’s Alpha and ITC must be interpreted with the items’ theoretical relevance to the measured construct.

EFA further reduced the number of questions to 26 (loaded into eight factors). However, two factors (each with two questions) were removed due to only two items loading into one factor. Therefore, the final questionnaire tested for fitness using CFA consisted of six factors and 22 questions.

### Limitation

This study was conducted from 2019 to 2020 during the COVID-19 pandemic. It received a study grant that required spending according to a strict timeline and progress reporting. The challenges started at the initial phase of the study when we were collecting inputs about the theoretical and response items from expert interviews. Discussions among experts were not as smooth as expected because of the absentees (panel contracting COVID-19), restriction of movement due to Movement Control Orders (MCO), or postponement of meetings [9]. Most of the interviews and discussions were conducted virtually. MCO was lifted by stages, but when the investigators were finally allowed to hire data collectors for EFA, the data collectors were only allowed to collect data within the hospital compounds with strict supervision from the hospital authorities. The inter-interviewer variation assessment could not be done before the EFA data collection because of the restricted exposure time allowed for each patient and the limited time allocation given to us to conduct repeated interviews. Test-retest interviews were also impossible as data collection was only allowed within the hospital compounds, and revisits at the subjects’ houses were not allowed.

The population in the country is diverse. Each region’s unique demographic, geographical profile, cultural differences, and practices may influence healthcare access and uptake. Cultural factors can influence how individuals interpret and respond to items on a scale, leading to factor loading and reliability variations across different cultural groups. In this study, the respondents in each region might have understood or interpreted the questions differently. Some questions might have different measurement scales for other cultures/regions, biases, or culture-specific, leading to different or low factor loadings and reliability [32]. The study should ideally be planned so that each region has a questionnaire tailored to the regional population profile (separate sets of development, validation and questionnaire data collection). This individual regional process and data collection in the community might result in a more robust EFA and reliability analysis (better factor loading, higher communality values, higher Cronbach Alpha value and better item reliability). However, a separate EFA would produce two questionnaires with possibly different domain/latent factors. A descriptive comparison between the regions would be difficult. Most importantly, the individual process would be more expensive and time-consuming (for this study and future studies). Adhering to the strict timeline and research grant protocol would be difficult.

The ‘inevitable’ flaw of the study was that data collectors were not allowed to collect data in the community due to the restriction of movement during the COVID-19 pandemic. This resulted in the responses being collected only from subjects (patients or relatives) in hospitals (mixed urban, district hospitals, and community health clinics). The responses should ideally be obtained from the community because the questionnaire was intended to identify influencing factors during the population survey at the community level. Obtaining responses from patients or relatives who might have preconceived ideas about knowledge, attitude and practice in cataract surgery probably skewed the results, leading to poor factor loading, low communalities and poor reliability of variables.

## Conclusion

This study validated a newly developed questionnaire using an EFA and reliability test. The questionnaire can potentially be used to identify the factors influencing cataract surgery uptake among individuals aged 50 years and above with VA worse than 6/18 (due to cataract) or individuals who have undergone cataract surgery. The final Questionnaire after EFA consisted of 6 factors with 22 items. All items demonstrated acceptable psychometric properties and had moderate reliability results.

## Data Availability

All survey data are available from the database URL https://www.raab.world/survey-data

## Acknowledgements

The authors would like to thank the Director General of the Ministry of Health Malaysia for his kind permission to publish this article. The authors would like to acknowledge the contribution of data collection and entry by the MyStep Personnel in making the EFA possible.

## Funding

Malaysian Ministry of Health grant

## Competing interests

None

## Ethics approval

Ethical approval was obtained from the Medical Research and Ethics Committee of the Malaysian Ministry of Health (Research ID NMRR-19-197-46172). The study was conducted in accordance with the tenets of the Declaration of Helsinki.

## Notes

### Competing Interest Statement

The authors have declared no competing interest.

### Author Declarations

Medical Research and Ethics Committee of the Malaysian Ministry of Health (Research ID NMRR-19-197-46172).

